# A phenome-wide association study of four syndromic genes reveals pleiotropic effects of common and rare variants in the general population

**DOI:** 10.1101/19010736

**Authors:** Catherine Tcheandjieu, Matthew Aguirre, Stefan Gustafsson, Priyanka Saha, Praneetha Potiny, Melissa Haendel, Erik Ingelsson, Manuel A. Rivas, James R. Priest

## Abstract

The clinical evaluation of a genetic syndrome relies upon recognition of a characteristic pattern of signs or symptoms to guide targeted genetic testing for confirmation of the diagnosis. However, individuals displaying a few phenotypes of a complex syndrome may not meet criteria for clinical diagnosis or genetic testing. Here, we present a phenome-wide association study (PheWAS) approach to systematically explore pleiotropy of common and rare alleles in genes associated with four well-described syndromic diseases (Alagille (AS), Marfan (MS), DiGeorge (DS), and Noonan (NS) syndromes) in the general population.

Using human phenotype ontology (HPO) terms, we systematically mapped 60 phenotypes related to AS, MS, DS and NS in 337,198 unrelated white British from the UK Biobank (UKBB) based on their hospital admission records, self-administrated questionnaires, and physiological measurements. We performed logistic regression adjusting for age, sex, and the first 5 genetic principal components, for each phenotype and each variant in the target genes (*JAG1, TBX1, FBN1, PTPN11, NOTCH2*, and *MAP2K1*) and performed a gene burden testing.

Overall, we observed multiple phenotype-genotype correlations, such as the association between variation in *JAG1, FBN1, PTPN11* and *SOS2* with diastolic and systolic blood pressure; and pleiotropy among multiple variants in syndromic genes. For example, rs11066309 in *PTPN11* was significantly associated with a lower body mass index, an increased risk of hypothyroidism and a smaller size for gestational age, all in concordance with NS-related phenotypes. Similarly, rs589668 in *FBN1* was associated with an increase in body height and blood pressure, and a reduced body fat percentage as observed in Marfan syndrome.

Our findings suggest that the spectrum of associations of common and rare variants in genes involved in syndromic diseases can be extended to individual phenotypes within the general population.

**Author Summary:** Standard medical evaluation of genetic syndromes relies upon recognizing a characteristic pattern of signs or symptoms to guide targeted genetic testing for confirmation of the diagnosis. This may lead to missing diagnoses in patients with silent or a low expressed form of the syndrome. Here we take advantage of a rich electronic health record, various phenotypic measurements, and genetic information in 337,198 unrelated white British from the UKBB, to study the relation between single syndromic disease phenotypes and genes related to syndromic disease. We show multiple phenotype-genotypes associations in concordance with phenotypes variations found in syndromic diseases. For example, we show that mutation in *FBN1* was associated with high standing/sitting height ratio and reduced body fat percentage as observed in individuals with Marfan syndrome. Our findings suggest that common and rare alleles in SD genes are causative of individual component phenotypes present in a general population; further research is needed to characterize the pleiotropic effect of alleles in syndromic genes in persons without the syndromic disease.

## Introduction

Genetic syndromes are rare diseases defined by a specific and clinically recognizable set of phenotypes across multiple organ systems. The era of next-generation sequencing has enabled substantial progress in linking syndromic disease to specific genetic loci, coupled with public databases of genotype-phenotype relationships to facilitate the classification of genetic variants from “benign” to “pathogenic” for use in clinical decision making. Large population-scale databases of genetic variation without phenotypes, such as ExAC, have provided additional context for characterizing genotype-phenotype relationships in genetic disease[1]. For mutations previously thought to cause disease, population databases have often suggested lower estimates of penetrance than initially recognized[2,3].

The diagnosis or classification of an individual with genetic syndrome relies upon expert recognition of a characteristic pattern of signs or symptoms or a set of defined diagnostic criteria. However, individuals displaying single phenotypes of a complex syndrome may not meet criteria for clinical diagnosis or genetic testing; expanding a binary definition of syndromic phenotypes to phenotype scores can identify more individuals with Mendelian disease patterns[4]. Similarly, individuals with clearly pathogenic mutations may be affected with only a single component phenotype of a genetic syndrome[5,6]. Recent descriptions of allelic heterogeneity, penetrance, and expressivity in syndromic disease genes have focused almost exclusively upon rare or familial alleles[7,8].

Here, we present a phenome-wide association study (PheWAS) approach to systematically explore pleiotropy of common and rare alleles in genes associated with four well-described syndromic diseases in the general population. Using the UK Biobank, we linked individual-level medical and morphometric data to the characteristic phenotypes of Alagille (AS), Marfan (MS), DiGeorge (DS), and Noonan (NS) syndromes. These data allow a survey of the association of common and rare alleles to single component phenotypes of each syndrome within the general (non-syndromic) population.

## Results

Based on the Human Phenotype Ontology (HPO) – an ontology-based system developed using medical literature and other ontology-based systems[9] – we identified 196 HPO terms related to AS, MF, DS, and NS. Of these 196 HPO terms, 53 were shared between at least two syndromes, and seven terms were included in all four syndromes (S1 Table). After grouping the HPO terms into categories based on affected organ systems, there were 115 HPO terms of which 73 could be matched to 100 phenotypes available in the UKBB. Most of the unmatched phenotypes were related to specific abnormalities of body structure or the musculoskeletal system, which were poorly represented in clinical and billing codes, or measurements such as impaired T-cell function, not available in the UKBB.

### Characteristics of the study population

A total of 337,198 unrelated individuals were included in our analysis; the mean age was 65.8 years (sd=8.0) and 53.7% of subjects were male. The number of subjects by phenotype is present in the S2 Table. Hypercholesterolemia (HP0003124), gastroesophageal reflux (HP0002020), premature osteoarthritis (HP0003088), and hypertriglyceridemia (HP0002155) were the most prevalent phenotypes with 12.8% (43,054 cases), 9% (30,229 cases), 8.9% (2,994 cases), and 8.6% (29,137 cases), respectively.

### Genotype-phenotype associations are common across syndromic genes

Overall, 9 phenotypes: hypothyroidism (HP0000821), diastolic BP (HP0005117), systolic BP (HP0004421), standing/sitting height ratio (abnormality of body height; HP0000002), birth weight (small for gestational age; HP0001518), amount of subcutaneous adipose tissues or body fat percent (reduced subcutaneous adipose tissue; HP0003758), growth abnormality (HP0001507), body mass index (abnormality of body mass; HP0045081), and hyperlipidemia (HP0003124) were significantly associated with multiple SNPs across *PTPN11, FBN1, JAG1, SOS2, RIT1, RAF1, KAT6B, RASA2, MAP2K1, DGCR2 and COMT* (Figure 1a, Table 1). The top SNP-phenotype association in each gene are reported in Table 1. Diastolic BP and systolic BP along with body mass index displayed a genetic association in each of the four syndromes. Birth weight, subcutaneous adipose tissue (body fat percent), and tall stature for MS or short stature for NS are common phenotypes for MS and NS; while hypothyroidism, and growth retardation are reported in NS and DS. After correction for multiple testing, several other phenotypes reach significance at the gene level (Figure 1b). When assessing the association by group of phenotypes in each syndrome, we observed different patterns of association.

**Table 1:** Table of association between the top SNPs in each gene and significant (pvalue < 3.16×10^−07^) or suggestive (p-value < 5×10^−04^) associations with HPO phenotypes.

| SD genes | Phenotypes (HPO code) | N Cases | N controls | Total | OR or BETA | Se or IC [95%] | P |
| --- | --- | --- | --- | --- | --- | --- | --- |
| MS | **15:48835935_C_T (rs589668); *FBN1* ALT=T Freq=0.25** |  |  |  |  |  |  |
|  | Standing and sitting height ratio (HP0000002) | - | - | 334227 | 0.030 | 0.0028 | 3.54E-27 |
|  | Diastolic blood pressure (HP0005117) | - | - | 334777 | 0.016 | 0.0028 | 7.00E-09 |
|  | Subcutaneous adipose tissue (HP0003758) | - | - | 331110 | -0.074 | 0.0179 | 3.52E-05 |
| NS | **12:112883476_G_A (rs11066309); *PTPN11* ALT=A Freq=0.41** |  |  |  |  |  |  |
|  | Hypothyroidism (HP0000821) | 17746 | 256695 | 337198 | 1.190 | [1.16-1.21] | 6.62E-59 |
|  | Diastolic blood pressure (HP0005117) | - | - | 334777 | 0.028 | 0.002 | 6.71E-30 |
|  | Standing and sitting height ratio (HP0000002) | - | - | 334227 | 0.016 | 0.002 | 4.59E-11 |
|  | Small for gestational age (HP0001518) | - | - | 192869 | -0.020 | 0.003 | 2.95E-10 |
|  | Systolic blood pressure (HP0004421) | - | - | 334544 | 0.014 | 0.002 | 3.24E-09 |
|  | Body mass index (HP0045081) | - | - | 333117 | -0.012 | 0.002 | 1.13E-06 |
|  | Stroke (HP0001297) | 4930 | 269511 | 337198 | 1.080 | [1.04-1.12] | 2.96E-05 |
|  | **14:50655357_G_C (rs72681869); *SOS2* ALT=C Freq=0.01** |  |  |  |  |  |  |
|  | Diastolic blood pressure (HP0005117) | - | - | 334777 | -0.076 | 0.0115 | 5.42E-11 |
|  | Systolic blood pressure (HP0004421) | - | - | 334544 | -0.070 | 0.0110 | 1.58E-10 |
|  | Subcutaneous adipose tissue (HP0003758) | - | - | 331110 | -0.452 | 0.0739 | 9.83E-10 |
|  | Body mass index (HP0045081) | - | - | 333117 | -0.055 | 0.0116 | 2.00E-06 |
|  | Small for gestational age (HP0001518) | - | - | 192869 | 0.072 | 0.0152 | 2.03E-06 |
|  | Inguinal hernia (HP0000023) | 367 | 7068 | 337198 | 1.230 | [1.11-1.38] | 1.61E-04 |
|  | Growth abnormality (HP0001507) | - | - | 328546 | -0.092 | 0.0257 | 3.61E-04 |
|  | **15:66702345_C_A (rs56913458); MAP2K1 ALT=A Freq=0.23** |  |  |  |  |  |  |
|  | Subcutaneous adipose tissue (HP0003758) | - | - | 331110 | -0.106 | 0.0185 | 1.14E-08 |
|  | Body mass index (HP0045081) | - | - | 333117 | -0.016 | 0.003 | 2.05E-08 |
|  | **1:155865042_T_C (rs61813631); RIT1 ALT=C Freq=0.02** |  |  |  |  |  |  |
|  | Small for gestational age (HP0001518) | - | - | 192869 | 0.067 | 0.013 | 2.32E-07 |
|  | **3:12649576_A_G (rs2442812); RAF1 ALT=G Freq=0.45** |  |  |  |  |  |  |
|  | Standing and sitting height ratio (HP0000002) | - | - | 334227 | 0.013 | 0.002 | 1.83E-08 |
|  | Hypercholesterolemia (HP0003124) | 37775 | 262539 | 337198 | 0.970 | [0.95-0.98] | 2.46E-05 |
|  | Intellectual disability, mild (HP0001256) | - | - | 108818 | -0.016 | 0.004 | 1.50E-04 |
|  | **3:141301451_G_A (rs3821710); RASA2 ALT=A Freq=0.40** |  |  |  |  |  |  |
|  | Growth abnormality (HP0001507) | - | - | 328546 | 0.036 | 0.006 | 4.80E-11 |
|  | Subcutaneous adipose tissue (HP0003758) | - | - | 331110 | 0.099 | 0.016 | 3.29E-10 |
|  | Body mass index (HP0045081) | - | - | 333117 | 0.010 | 0.002 | 8.33E-05 |
|  | Body height (HP0000002) | - | - | 334227 | -0.009 | 0.002 | 1.99E-04 |
|  | **10:76617857_C_T (rs12254441); KAT6B ALT=T Freq=0.37** |  |  |  |  |  |  |
|  | Subcutaneous adipose tissue (HP0003758) | - | - | 331110 | -0.096 | 0.017 | 7.12E-09 |
|  | Body mass index (HP0045081) | - | - | 333117 | -0.013 | 0.003 | 6.42E-07 |
|  | Hypothyroidism (HP0000821) | 10387 | 172953 | 337198 | 0.940 | [0.92-0.96] | 1.07E-07 |
| AS | **20:10624190_A_G (rs889509); JAG1 ALT=G Freq=0.42** |  |  |  |  |  |  |
|  | Diastolic blood pressure (HP0005117) | - | - | 334777 | -0.021 | 0.002 | 5.55E-17 |
|  | Small for gestational age (HP0001518) | - | - | 192869 | -0.012 | 0.003 | 1.86E-04 |
| DS | **22:19104926_C_G (rs807747); DGCR2 ALT=G Freq=0.06** |  |  |  |  |  |  |
|  | Body height (HP0000002) | - | - | 334227 | -0.028 | 0.005 | 1.16E-08 |
|  | **22:19960184_C_G (rs71313931); COMT ALT=G Freq=0.29** |  |  |  |  |  |  |
|  | Systolic blood pressure (HP0004421) | - | - | 334544 | 0.013 | 0.003 | 1.88E-07 |
|  | Body mass index (HP0045081) | - | - | 333117 | 0.010 | 0.003 | 3.38E-04 |

**Table 2:** table summarizing number of genes, SNPs and phenotypes (HPO terms) for each syndrome, include in our analysis

|  | Genes | MAF>0.005,<br>R2<0.8 | HPO<br>terms | matched<br>HPO term |
| --- | --- | --- | --- | --- |
| Alagille Syndrome | *JAG1, NOTCH2* | 302 | 61 | 36 |
| Marfan syndrome | *FBN1* | 518 | 64 | 42 |
| Noonan syndrome | *PTPN11, SOS1, RAF1, KRAS, RIT1, BRAF, A2ML1, RRAS, SOS2, NRAS, RASA2, CBL, SHOC2, MAP2K1, KAT6B* | 3062 | 58 | 24 |
| DiGeorge syndrome | *22q11.2 deletion (TBX1, DGCR2, DGCR8, DGCR6, COMT, PRODH, HIC2, LZTR1)* | 1228 | 66 | 35 |

**Figure 1.**
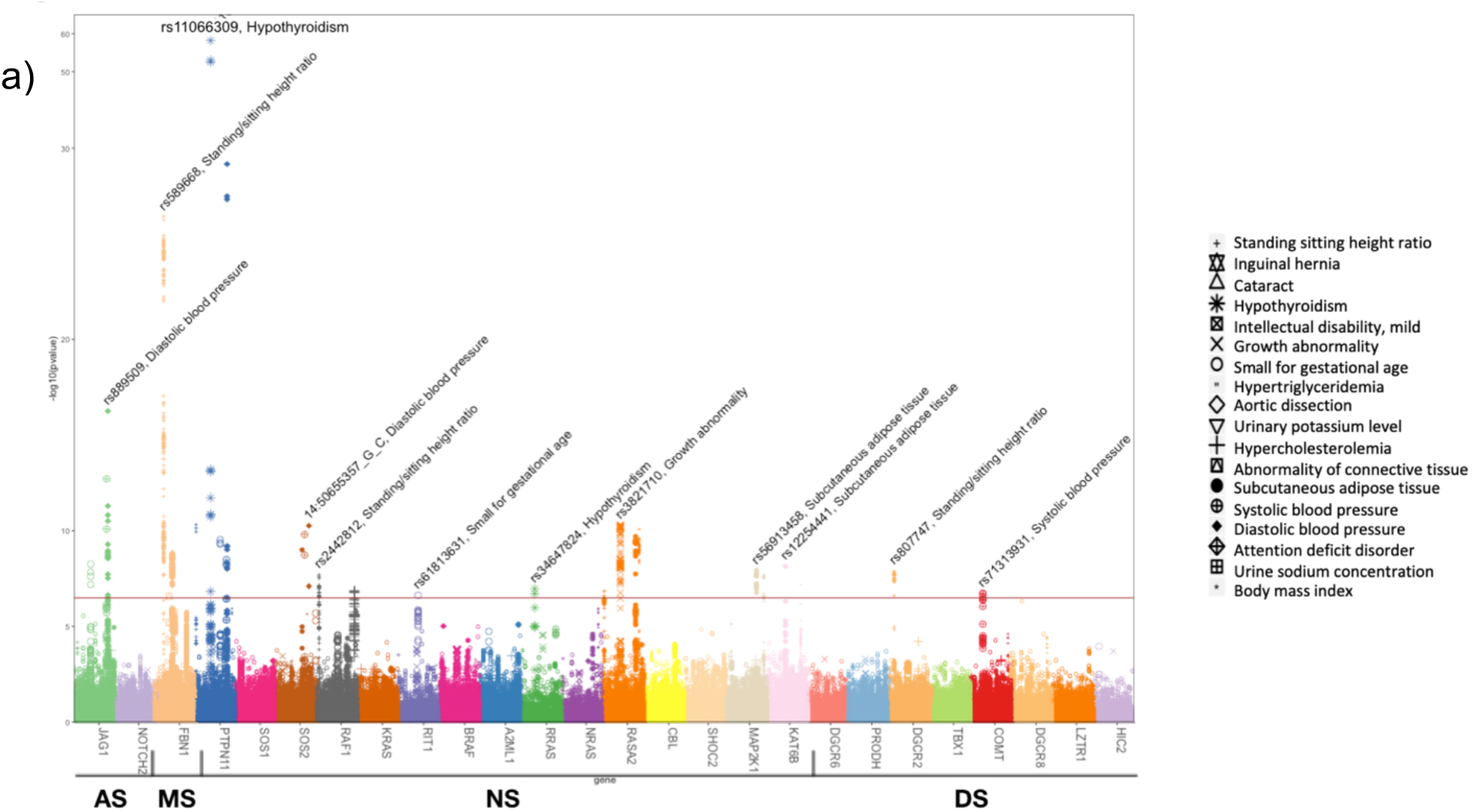
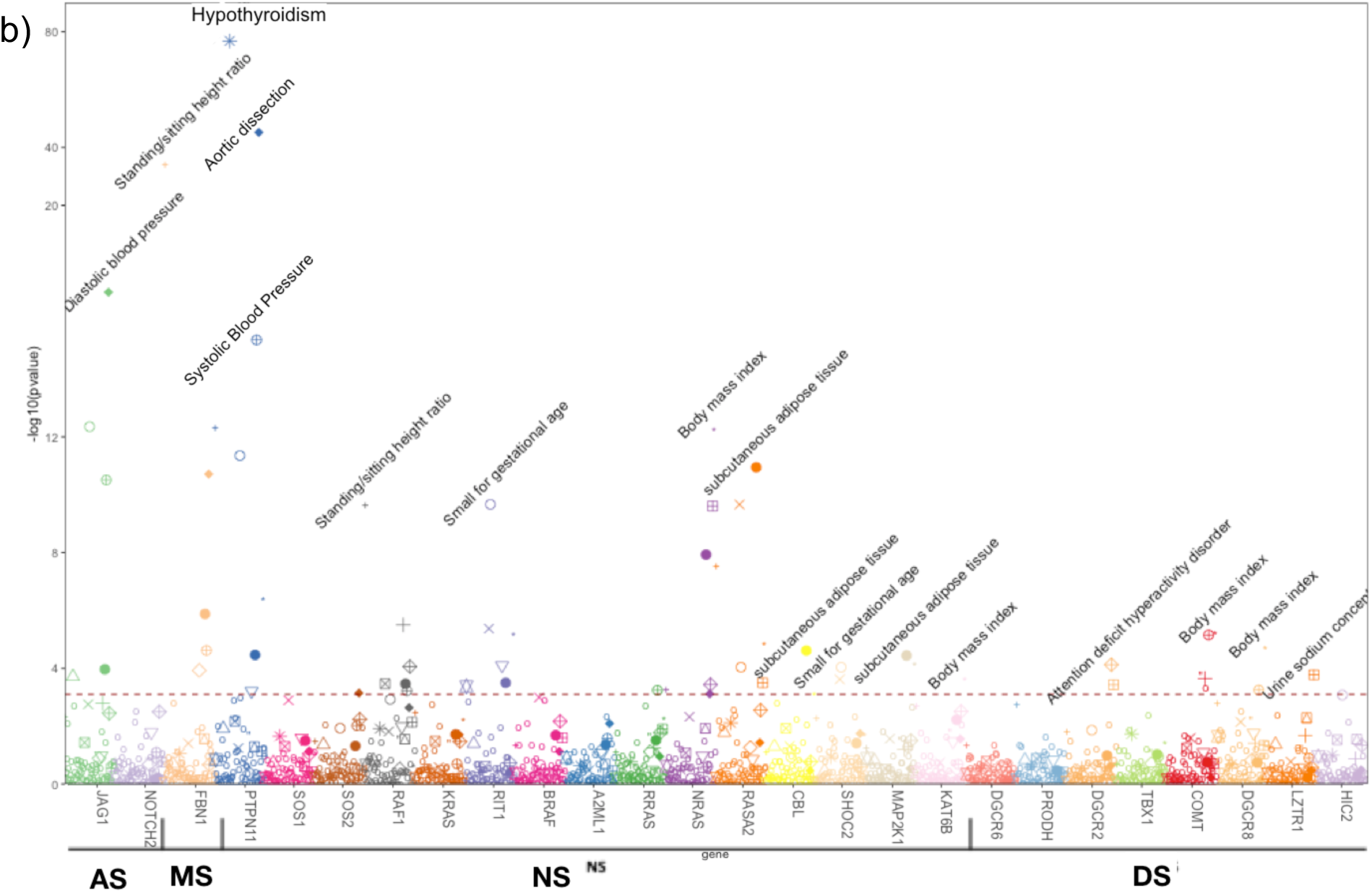
Primary PheWAS results: the variant level (a) and gene level (b). The red line represents the level of significance after Bonferroni correction (p<3.2×10^−07^ at SNP level and p<9×10^−04^ at gene level). Genes are represented by color and HPO terms are indicated by shape.

### Variation in syndromic genes are associated with component phenotypes

Marfan syndrome (MS) is a primary disorder of connective tissue with diagnostic criteria centered around cardiovascular, musculoskeletal, and ocular phenotypes linked to a single gene *FBN1* which encodes an extracellular matrix protein. Several SNPs in *FBN1* were significantly associated with increased standing/sitting height ratio and an elevated diastolic BP. An increased risk of aortic dissection and a lower percent of body fat (two major phenotypes in MS) were observed for several of these SNPs although the association was suggestive (Figure 2a and S3 Table). All SNPs in *FBN1* displaying associations were located within the same LD block and were highly correlated suggesting a single signal within the gene (Figure 2a and 2b). A common intronic variant rs589668 displays the top signal with high standing/sitting height ratio (beta=0.03, se=0.002, p=10^−27^, Table 1), an elevated diastolic BP (beta=0.02, se=0.002, p=7×10^−09^), and a lower percent of body fat (beta=-0.07, se=0,02, p=5×10^−05^, Table 1). Four additional SNPs (rs11070641, rs4775760, rs363832 and rs140605) that reach genome-wide significance with high standing/sitting height ratio and diastolic BP were correlated with several syndromic disease entities in CLINVAR including stiff skin syndrome, ectopia lentis, MASS syndrome, thoracic aortic aneurysm and aortic dissection (Supplementary Table 3). At the gene level association using SKAT test with SNPs allelic frequency weighted by their CADD (Combined Annotation Depletion Dependent) score, standing/sitting height ratio, systolic and diastolic BP, subcutaneous adipose tissue, and aortic dissection were significantly associated with *FBN1* (Figure 1b, S4 table).

**Figure 2:**
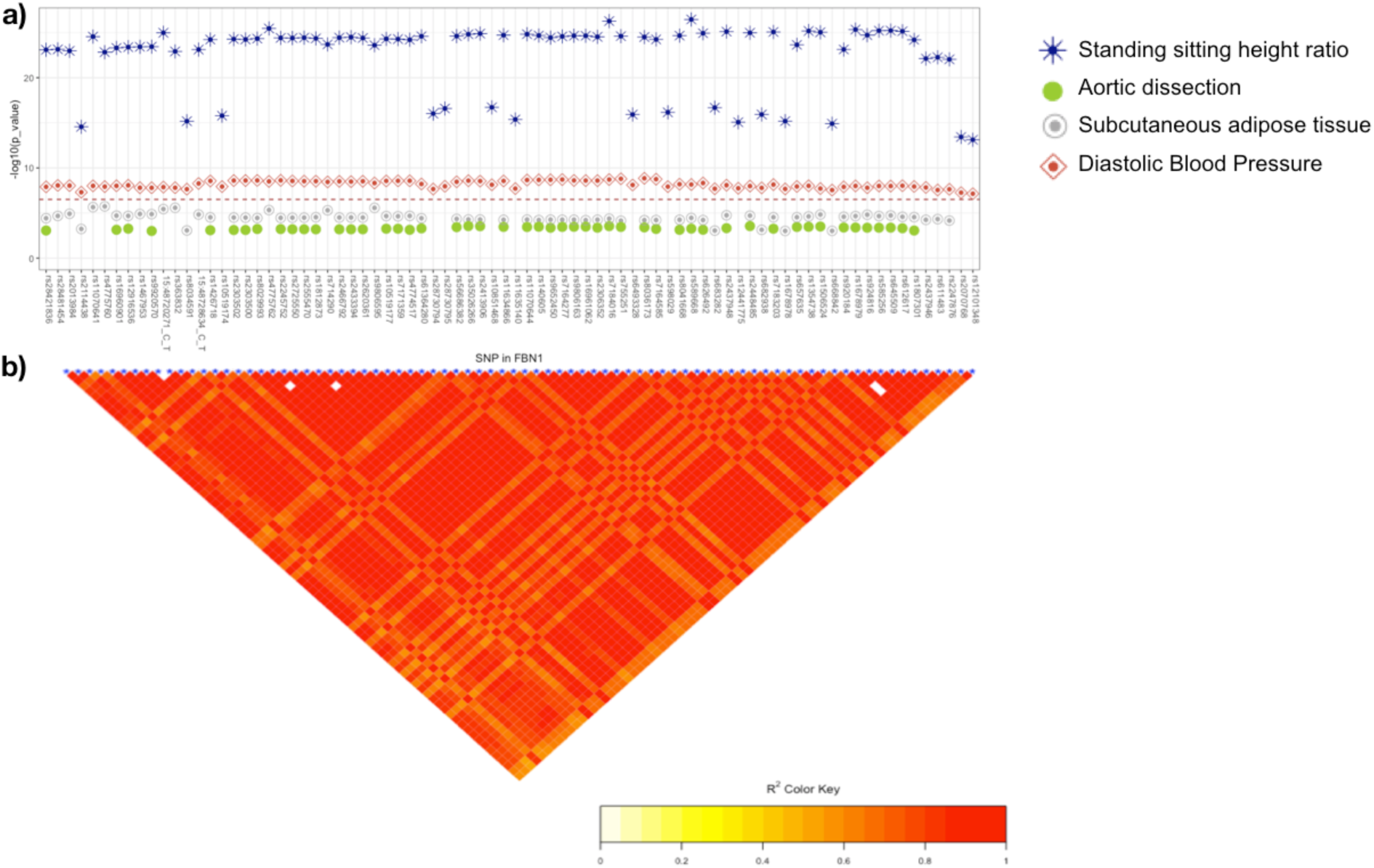
PheWAS result and linkage plot for SNPs with pleiotropy in *FBN1*. Associations between SNPs with Pleiotropy and phenotypes (a) and linkage between the SNPs in *FBN1* (b). The red line represents the level of significance after Bonferroni correction (p=3.2×10^−07^)

For NS and RAS-opathy related phenotypes, SNPs in *PTPN11* were associated with increased risk of hypothyroidism, high diastolic and systolic BP, and high standing/sitting height ratio (Figure 1a, S3 Table). SNPs in *SOS2* were associated with lower systolic and diastolic BP, and lower percent of body fat (Figure 1a, Supplementary Table 3). SNPs in *MAP2K1* were associated with lower body mass index, as well as lower level of cutaneous adipocytes tissues (Figure 1a, Supplementary Table 3). SNPs in *PTPN11* display a moderate to low correlation with each other suggesting several independent signals within the locus, while high correlation was observed between SNPs in *RASA2, SOS2* and *MAP2K1* indicating that the association observed within each gene represents a single signal (Figure 3b). Among SNPs with pleiotropic effects, rs11066309 in *PTPN11* displays a strong association with increased risk for hypothyroidism (ALT freq=0.40; OR [95% CI]: 1.19; [1.16 – 1.21]; p=6×10^−59^) along with five other phenotypes, including decreased body mass index (beta=-0.012, p=1.13×10^−06^) and birth weight (beta=-0.020, p=2.95×10^−10^) (Figure 3a, Table 1). In addition, among SNPs that reached significance, rs3741983 (*PTPN11*), rs72681869 (*SOS2*), rs61755579 (*SOS2*), and rs112542693 (*MAP2K2*) were linked to Noonan syndrome and a number of abnormalities including global developmental delay, short stature and abnormality of cardiovascular system in ClinVar. At a gene level, *PTPN11, NRAS, RASA2, SOS2, MAP2K1*, and *RAF1* were significantly associated with hypothyroidism, diastolic and systolic BP, birth weight, growth abnormality, subcutaneous adipose tissue, standing/sitting height ratio and body mass index (Figure 1b and S4 Table).

**Figure 3:**
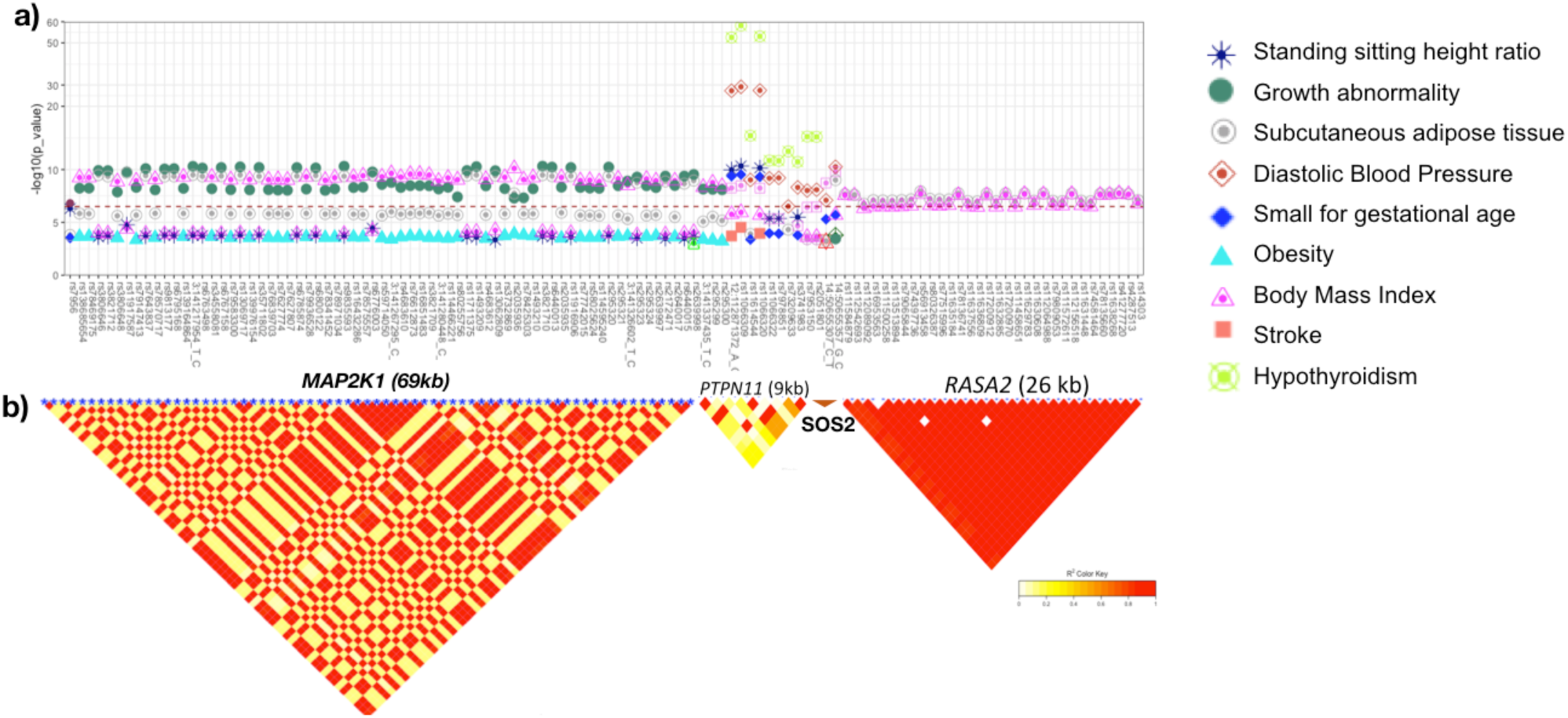
PheWAS result and linkage plot for SNPs with pleiotropy in RAS-opathie genes. Associations between SNPs with Pleiotropy effect in *MAP2K1, PTPN11, SOS2* and *RASA2*, and HPO terms (a) and linkage between the SNPs (b). The red line represents the level of significance after Bonferroni correction (p=3.2×10^−07^)

Alagille syndrome is caused by mutations in *JAG1* and *NOTCH2* with major clinical manifestations in the heart and liver, and characteristic facial features. At a SNP level, none of the AS specific phenotypes reached significance after multiple testing correction. However, suggestive associations were observed between rs1051412 (reported in ClinVar as a benign variant) and cataracts (p=2.9×10^−04^, S1(a) Figure). Several SNPs in *JAG1* were significantly associated with diastolic BP, systolic BP and birth weight (p<10^−08^, Figure 1a, S3 Table). The SNP rs889509 (20:10658882_G_C) displayed the most significant association with a lower diastolic BP (beta=-0.028; p=8.2×10^−11^, Table 1). At a gene level, *JAG1* was associated with diastolic BP (p=3.48×10^−15^), birth weight (p=3.84×10^−10^), systolic BP (p=3.32 ×10^−09^) and cataracts (p=2.21×10^−04^, Figure 1b, Supplementary Table 4).

DiGeorge syndrome encompasses a recurrent microdeletion of multiple genes at the 22q11.2 locus due to the presence of segmental duplications, with affected individuals displaying neuropsychiatric, immunological, and cardiovascular phenotypes originating from defects in neural crest cell formation and migration. At the SNP level, rs807747 in *DGCR2* and rs71313931 in *COMT* were significantly associated with abnormal body height (p=2×10^−07^) and systolic BP (p=5.6×10^−17^), respectively (Figure 1a and Table 1). At the gene level, *COMT* was associated with systolic BP, while *DGCR2* was associated with diastolic BP and standing/sitting height ratio (p<5×10^−4^) (S4 Table).

## Discussion

Here, we systematically describe the association of variation at four syndromic loci with the component phenotypes of syndromic disease. We hypothesize that in the general population, common and rare alleles for syndromic diseases display pleiotropic effects with the phenotypes related to genetic syndromes. Using the UKBB, we linked individual-level data to the characteristic phenotypes of Alagille, Marfan, Noonan, and DiGeorge syndromes, showing clearly the association of common and rare alleles to single component phenotypes of each syndrome. While classical Mendelian forms of each syndrome may be uncommon, individual phenotypes attributable to these syndromic loci appear to be common within the general population. This finding support the findings of Bastarache et al[4], suggesting a scaling of Mendelian disease phenotypes into a continuous phenotyping score to improve the identification of individuals with rare diseases.

Within families of individuals affected by syndromic disease carrying the same pathogenic mutation, the expressivity of component phenotypes may vary in different individuals[10,11]. Here, we show that many common and rare variants at syndromic loci existing in the general population may result in expression of traits and phenotypes closely related to the syndrome of interest. For example, we observe associations of a common intronic variant in *FBN1* rs589668 (MAF=0.25 in Europeans populations) with increases in blood pressure and height and decreased subcutaneous fat distribution. In GTex[12], this variant is an eQTL strongly associated with decreased expression in whole blood (p=1.7×10^−37^), which would be concordant with the known molecular mechanism of *FBN1* pathogenesis in MS: pathogenic alleles impairing gene function result in increased height and abnormal fat distribution and increased arterial stiffness[13,14]. Modifiers of penetrance and phenotypic expressivity in Marfan syndrome have been proposed,[15,16] but our results suggest that common variants and local haplotype structure around syndromic genes may deserve more attention[17].

Noonan syndrome is caused by mutations in *PTPN11* and part of a group of related disorders arising from activating mutations in RAS-MAPK signaling pathway known as RASopathies which display many phenotypes across a variety of organ systems. A wide phenotypic variability and genetic heterogeneity have also been described in individuals with Noonan syndrome in relation to rare variants in *PTPN11*[18]. Here, we show that even in the general population, common and rare variants in *PTPN11* are independently associated with phenotypes such as hypothyroidism, small birth weight and low percent of body fat observed in some Noonan syndromes cases[19–21]. In GTex[12], numerous variants in *PTPN11*, such as rs11066309, rs3741983 and rs11066322 were significantly associated with a decreased expression in atrial appendage, adipose tissue, thyroid and skin and esophagus. Although in consistent with the role of *PTPN11* in thyroid function, cancer and autoimmunity[22–24], these variants are instead described as eQTLs with *TMEM116, ALDH2* and *MAPKAPK5-AS1* located up to 500kb upstream of *PTPN11*, suggesting that the association observed with rs11066309, rs3741983 and rs11066322 may also be potentially linked to other genes.

Growth retardation, lower BMI and short stature are well-known characteristics of Noonan syndrome, and a recent study reported a phenotype-genotype variability of growth pattern in patients with Noonan syndrome[25]. In concordance with this study, we showed that, in the general population, common and rare variants in *RASA2, SOS2* and *MAP2K1* are independently associated with growth characteristics (body mass index, height and growth abnormality) and the association driven by single or multiple haplotype in each gene.

When performing genetic testing, allele frequency is often used as a marker to determine pathogenicity of a genetic variant. Common variation in and around *JAG1* has previously been associated with such disparate phenotypes as pulse pressure, circulating blood indices, and birthweight, and none of the variants included in our analysis appeared to be directly associated with the component phenotypes of AS. However, the unifying molecular abnormality in AS are defects in vascular formation which lead to each of the component cardiovascular and liver phenotypes of Alagille syndrome[26,27].

Our study has some limitations. Our analysis was limited to phenotypes with more than 100 cases; therefore, diseases with relatively rare prevalence were not analyzed. In addition, because our study cohort consist of adults from the general population, specific phenotype targeting facial and skeletal dysmorphism, such as butterfly vertebrae or broad forehead; specific abnormalities of organs, such as biliary disease; and phenotypes observed during childhood, such as developmental delay or attention deficit and hyperactivity disorder (ADHD) were not present. However, to correct on the lack of some of these phenotypes, we used certain phenotypes or measurement present in the UKBB, such as head circumference as a proxy for broad forehead, education level for ADHD, weight and height at age 10 as proxy for growth abnormality. That said, the complexity of matching UKBB phenotypes to HPO terms may simply not capture some phenotypes, despite manual curation.

Key strengths of our study include the ability to systematically test multiple phenotype-genotype association and to highlight pleiotropy linked to syndromic genes. Our study maps UKBB phenotypes to HPO terms and shows that common and rare variants in genes responsible for Alagille, Marfan, Noonan, and DiGeorge syndromes, are also independently associated with component phenotypes of these syndromes in the general population.

Our findings suggest that the spectrum of association of common and rare variants in genes involved in syndromic diseases can be extended to individual component phenotypes in the general population. Further research is needed to characterize the pleiotropic effect of alleles in genes in persons without the corresponding genetic syndrome.

## Materials and Methods

### Study population and data collection

The study cohort was derived from the UK Biobank (UKBB), a large prospective cohort study with comprehensive health data from over 500,000 volunteer participants in the United Kingdom aged 37-73 years at recruitment in 2006-2010. The cohort has previously been described in detail[28,29]. Information on the UK biobank participants was collected at enrollment, and from electronic health record (EHR) information which includes diagnostic codes (ICD10, ICD9) and procedural code (OPCS) from hospital admission records dating to 1992, and cancer registries. Data collected at the assessment visit included information on a participant’s health and lifestyle, hearing and cognitive function, collected through a touchscreen questionnaire and verbal interview. A range of physical measurements was also performed, including blood pressure; arterial stiffness; body composition measures (including impedance); hand-grip strength; ultrasound bone densitometry; spirometry; and an exercise/fitness test with ECG. Samples of blood, urine, and saliva were also collected. Medical phenotypes were aggregated as previously described, incorporating available information including a broad set of medical phenotypes defined using computational matching and manual curation of on hospital in-patient record data (ICD10 and ICD9 codes), self-reported verbal questionnaire data, and cancer and death registry data[30,31].

### Phenotypes of target syndromes

We identified phenotypes related to syndromic diseases through the Human Phenotype Ontology (HPO). HPO is an ontology-based system developed using medical literature, and other ontology-based systems such as Orphanet, and OMIM[9]. HPO provides a standardized vocabulary of phenotypic and abnormalities encountered in human diseases. The HPO has link symptoms/phenotypes to diseases or genetic disorders, and the causing genes. As an example, Alagille syndrome (AS) is linked to *JAG1*, and *NOTCH2* genes as well as all the phenotypes or symptoms observed in AS, such as atrial septal defect, hypertelorism, and butterfly vertebra.

HPO terms were directly matched to UKBB phenotypes when phenotypes in both systems had similar terminology. The direct phenotype matching was conducted using a semi-automatic mapping system which combines semantic and lexical similarity between word[32] followed by manual curation. When the HPO terms were not present, we performed an indirect matching by hand to find in the UKBB, the phenotype that best reflects the target HPO terms. For example, abnormality of body structure or body morphology such as abnormal body height, reduced sub cutaneous adipose tissues, bone density or broad forehead, were respectively matched to sitting/standing height ratio; body fat percentage; bone mineral density, and head bone area. For psychiatric diseases such as depression and neurodevelopmental disorders such as attention deficit and hyperactivity disorder (ADHD), we used a score of depressive symptoms and self-reported educational level respectively as proxies for these terms.

To increase the number of subjects in some subgroup of phenotypes, we combined subcategories of HPO terms into a group or category. For example, 39 HPO terms representing an abnormality of head, ears, and eyes such as low-set ears, strabismus, macrotia, webbed neck, short neck, abnormality of the eye, microcornea, down-slanted palpebral fissure and other congenital abnormality of ears, were grouped into “Abnormality of head or neck (HP0000152)” and mapped to icd10 targeting congenital malformations of eye, ear, face, and neck and other organs especially facial appearance (ICD10: Q10 to Q18 and Q87). Ten HPO terms for congenital abnormality of cardiovascular system including Ventricular septal defect, Atrial septal defect, Tetralogy of Fallot, Patent ductus arteriosus, Bicuspid aortic valve, Truncus arteriosus, Coarctation of aorta, Tricuspid valve prolapse were combined into abnormality of the cardiovascular system (HP0001626) and mapped to Congenital malformations of the circulatory system (ICD10: Q20 to Q28).

### Genotyping data

Genotyping was performed using the Affymetrix UK BiLEVE Axiom array on an initial 50,000 participants; the remaining 450,000 participants were genotyped using the Affymetrix UK Biobank Axiom® array. The two arrays are extremely similar (with over 95% common content). Quality control and imputation to over 90 million SNPs, indels and large structural variants was performed[33]. 5647 SNPs in 26 genes related to Alagille syndrome, Marfan syndrome, Noonan syndrome, and DiGeorge syndrome were selected for our study (table2). The selected SNPs had a MAF>0.005 and an imputation measurement (R2) ≥ 0.8

### Statistical analysis

#### SNP level

For binary traits, logistic regression with adjustment on age, sex, batch, and the top 5 principal components were used. First, regression was used in a situation of unbalanced numbers of cases and controls, especially when the number of cases was very small (less than 200 cases). For continuous traits, we performed linear regression with adjustment on age, sex, batch, and the top 5 principal components. Our analysis was restricted to individuals of Europeans descent, due to the relatively small number of individuals from other ethnic groups in the UKBB. Bonferroni correction based on the number of independent tests was used to correct on multiple testing. Given the high correlation between SNPs within gene or regions, Bonferroni correction is often stringent when to number of tests considered is number of SNPS time the number of phenotypes. To take in account the correlation between SNPs, we estimate the number of independent SNPs in a block of 50 kb with a correlation > 0.8 using the pairwise pruning method implemented in <u>PLINK</u> which estimated 2166 independent SNPs within our target regions. We apply a threshold of 3.16×10^−07^=0.05/(2166×73) independent tests. Gene Level

The Sequence Kernel association test (SKAT) was performed for gene-level association using allelic frequency methods and weighted by the CADD score (Combined Annotation Depletion Dependent)[34,35]. A Bonferroni correction of 1×10^−03^ based on 26 genes analyzed using allelic frequency methods and weighted by the CADD score was applied for multiple testing correction.

## Data Availability

Additional results can be found in supplementary materials.

## Acknowledgments

This study is supported by data from the UKBB, applications 24983 (Dr Rivas), 15860 (Dr Priest), and 13721 (Dr Ingelsson).

## Supporting information

**S1 Table**: Overall HPO term present in Alagille, Noonan, Marfan and Digeorge syndromes

**S2 Table**: description binary and continuous phenotypes

**S3 table**: Association between all SNPs and all phenotypes

**S4 table**: Gene level association with SKAT with SNPs weighted using the CADD score or using the MAF

**S1 figure**: diagram phenotype matching system between the UK-biobank (UKBB) phenotypes and HPO terms

**S2 figure**: Association between SNPs in *FBN1* and all HPO terms at the SNPS and gene levels

**S3 figure**: Association between SNPs in *PTPN11*, and *gene in RAS/MAKP2* and all HPO terms at the SNPS and gene levels

**S4 figure**: Association between SNPs in *NOTCH2*, and *JAG1* and all HPO terms at the SNPS and gene levels

**S4 figure**: Association between SNPs in *22q11 locus* and all HPO terms at the SNPS and gene levels

**S6 figure**: Forest plots showing association between Clinvar SNPs in *FBN1, JAG1, PTPN11, MAP2K1* and *SOS2* and, MF, AS, NS-related phenotypes, respectively.

